# Persistent Fibromyalgia-like symptom presentation in Long COVID-associated chronic musculoskeletal pain: a longitudinal cohort study

**DOI:** 10.64898/2026.10.07.26364942

**Authors:** Bárbara Silva-Passadouro, Ioannis Delis, Alexander J. Casson, Christopher Brown, Sarah L. Astill, Manoj Sivan

**Affiliations:** Leeds Institute of Rheumatic and Musculoskeletal Medicine, School of Medicine, Faculty of Health and Life Sciences, University of Leeds, Leeds, UK; School of Biomedical Sciences, Faculty of Health and Life Sciences, University of Leeds, Leeds, UK; School of Electrical and Computer Engineering, National Technical University of Athens, Athens, Greece; Department of Electrical and Electronic Engineering, University of Manchester, Manchester, UK; Department of Psychology, Institute of Population Health, University of Liverpool, Liverpool, UK; National Demonstration Centre in Rehabilitation Medicine, Leeds Teaching Hospitals NHS Trust, Leeds, UK

**Keywords:** Post-COVID syndrome, Post Acute Covid Syndrome, Fibromyalgia Syndrome, Chronic pain, Neuropathic pain

## Abstract

**Background:** Chronic musculoskeletal pain (CMP) is a common and debilitating symptom of Long COVID (LC), which shares several clinical features with Fibromyalgia Syndrome (FMS).

**Aims:** This longitudinal study investigated whether the symptom overlap between LC-CMP and FMS persists over time, whether patterns differ according to pre-COVID CMP history, and the extent to which neuropathic pain features are present.

**Methods:** Individuals with LC completed questionnaires assessing the symptom-based component of the 2010 American College of Rheumatology criteria for FMS and the presence of neuropathic pain features (PainDetect Questionnaire, PDQ) at baseline, 6- and 12-month follow-ups. Participants were grouped according to pre-COVID CMP history. Group differences were analysed using mixed-effects logistic regression.

**Results:** 240 individuals completed the baseline assessment at a mean of 2.61 years after LC onset, including 131 (100 females) with new-onset CMP and 94 (87 females) with pre-existing CMP. The prevalence of participants meeting the symptom-based criteria for FMS remained high across follow-up (67.5–78.6%), with 50.0–61.0% of those with complete follow-up data fulfilling criteria at all timepoints. No significant subgroup differences were observed (OR = 0.43, 95% CI [0.14 1.39], p = 0.160). Across follow-up, 22.2–36.1% of participants reported likely neuropathic pain features, with significantly higher odds in the pre-existing CMP group (OR = 3.17, 95% CI [1.22 8.18], p = 0.017).

**Conclusions:** FMS-like symptoms remain highly prevalent several years after acute COVID-19 and persist over a further 12 months, suggesting that LC-CMP frequently represents a persistent long-term condition with substantial overlap with FMS. These findings support closer integration of LC and chronic pain rehabilitation services and add to the wider literature linking post-infectious processes with FMS-like symptoms.

## Introduction

Six years after the coronavirus disease 2019 (COVID-19) outbreak, its lasting effects continue to challenge healthcare systems worldwide. Many individuals developed and continue to experience persistent symptoms long after the initial Severe Acute Respiratory Syndrome Coronavirus 2 (SARS-CoV-2) infection, that constitute a novel clinical syndrome known as Long COVID (LC). Recent studies have reported that approximately 55–71% of individuals continue to experience at least one post-COVID symptom even two years after the acute infection, with chronic musculoskeletal (MSK) pain among the most commonly reported.^1–4^ Long COVID chronic musculoskeletal pain (LC-CMP) substantially impacts daily functioning and quality of life,^5,6^ adding to the global burden of chronic pain already affecting over 1.5 billion people.^7^ Despite this, LC-CMP remains poorly managed, likely reflecting lack of recognition and limited understanding of contributing mechanisms.

The widespread distribution of pain in LC-CMP and presence of comorbid symptoms of fatigue, sleep disturbances and cognitive impairments, resembles the clinical presentation of Fibromyalgia Syndrome (FMS). FMS is a complex multisystem condition characterised by widespread MSK pain that is not attributable to identifiable structural pathology in the MSK system.^8^ Although the precise aetiology of FMS remains unclear, there is growing evidence suggesting a role for viral infection as a potential trigger, in addition to other contributing factors including inflammation and autoimmunity.^9,10^ Increased rates of FMS have been demonstrated in individuals that contracted infection by Hepatitis C, HIV and the earlier coronavirus SARS-CoV, for instance.^11–13^ Regarding SARS-CoV-2, studies have shown that 14.2–31.0% of previously infected individuals fulfil the American College of Rheumatology (ACR) diagnostic criteria^14^ for FMS,^15–17^ with numbers going up to 72.2% among those with self-reported or physician diagnosed LC.^18–20^ This is substantially higher than the 2–4% prevalence of FMS reported in the general population.^21^

In addition to the considerable overlap in symptom profiles, LC-CMP and FMS exhibit similar demographic characteristics including a higher prevalence in females and associations with comorbid mood disorders.^1,22^ These similarities may reflect common susceptibility factors, multimorbidity, or broader post-viral processes, and have also led to the hypothesis of shared underlying pathophysiological mechanisms.^20,23,24^ FMS is primarily associated with nociplastic-type pain mechanisms, wherein sensitisation of the central and peripheral nervous systems contributes to altered nociception and pain amplification,^25^ which may similarly contribute to persistent pain in LC-CMP.^23^ The potential involvement of neuropathic pain mechanisms in some LC cases has also been hypothesised,^26^ with few studies reporting neuropathic-like sensory symptoms, such as burning, tingling and numbness, in a subset of individuals with LC-CMP.^27–29^ Identification of pain phenotypes in LC is clinically important, as it may help inform individualised therapeutic strategies and support more adequate and tailored pain management for these individuals.^30^

While previous studies have reported cross-sectional similarities between LC-CMP and FMS, little is known about the stability of this phenotype over time. Furthermore, it remains unclear whether individuals with LC pre-existing CMP differ phenotypically from those with LC new-onset CMP. Addressing these questions has important implications for clinical practice as it may help contextualise LC-CMP as a transient post-viral sequela or as a persistent chronic pain condition requiring long-term management approaches similar to those used for established chronic pain syndromes. The primary aims of this study were to investigate (a) whether the clinical overlap between LC-CMP and FMS remains evident several years after the acute COVID-19 infection and persists over a further 12-month period, and (b) whether this overlap differs according to the presence of pre-existing CMP. We also assessed the presence of neuropathic pain features in LC-CMP using a validated patient-reported screening tool.

## Methods

### Participants

Data presented here derive from the UNICO (Understanding Chronic Pain in Long COVID) study, a longitudinal questionnaire study that aimed to assess pain characteristics in individuals with LC, changes over time and associated factors. We recruited individuals self-reporting symptoms consistent with LC, persisting for at least 4 weeks following confirmed or suspected COVID-19 infection. Onset of symptoms must have occurred after December 2019. Further inclusion criteria included being 18 years or older, able to read and understand English, and residing in the UK. Participants were recruited primarily through promotion via social media and LC support groups, as well as through university mailing lists.

All participants provided digital informed consent through checkboxes, recorded digitally on the JISC Online Surveys platform (onlinesurveys.jisc.ac.uk, Bristol, UK), formerly Bristol Online Survey, prior to any data collection. It was made clear to the participant that by agreeing with the statements outlined and submitting their answers they consented for their data to be used for research. Participation in the study was entirely voluntary, and participants were informed of their right to withdraw at any time. Ethical approval was obtained from the University of Leeds’ Faculty of Medicine Ethics committee (MREC 22-019).

### Questionnaires

Patient-reported outcome measures were completed on the Digital Patient Reported Outcome Measure (DPROM) platform, developed by the digital health company ELAROS 24/7 Ltd and the University of Leeds.^31^ Participants were individually registered on the digital platform and accessed questionnaires through unique personal accounts, allowing responses to be attributed to individual participants. Where multiple responses to the same questionnaire were received from a participant at a given follow-up, only the first submitted response was included in the analysis. Demographic information and COVID-19 related history were collected at baseline (T0). Self-reported pain measures were assessed at T0 and scheduled for follow-up assessment at 6 months (T1) and 12 months (T2) after T0. The precise timing of completion of follow-up assessments varied between participants, and data were collected approximately around the scheduled follow-up date with median follow-up duration of 5.98 months (range: 5.55–7.36; IQR 5.91–6.08) for the 6-month follow-up (T0 to T1) and 11.96 months (range: 10.32–14.88; IQR 11.89–12.07) for the 12-month follow-up (T0 to T2). Participants who had not completed the survey by the scheduled date received up to four email reminders to help maximise response rates. Those who did not complete the T1 follow-up assessment were still invited to complete the T2 follow-up.

The severity of common symptoms of LC was assessed using the Modified COVID-19 Yorkshire Rehabilitation Scale (C19-YRSm).^32^ The scale includes pain items that ask individuals to rate their pain from 0 = no problem to 3 = severe problem, relative to symptoms before (pre-COVID) and after contracting LC (post-COVID). Participants were considered to present with MSK pain if reporting scores greater than zero on the joint and/or muscle pain items.

Fulfilment of the 2010 American College of Rheumatology (ACR) criteria for FMS was examined based on the Widespread Pain Index (WPI) and Symptom Severity (SS) scales.^14^ The WPI is a checklist of 19 body areas that assesses the distribution of pain (range 0–19). The SS scale measures the severity of fatigue, waking unrefreshed and cognitive symptoms (0 = no problem, 3 = severe problem), as well as the presence of other somatic symptoms (score of 0–3 depending on the number of symptoms selected from a list of 40). The scores are combined to assess whether the participant presents with an FMS-like phenotype based on the symptom-based component of the 2010 ACR criteria: WPI score ≥ 7 and SS score ≥ 5, or WPI score between 3–6 and SS score ≥ 9. Due to the questionnaire-based design of the study, the requirement that symptoms are not explained by an alternative diagnosis, could not be independently verified and would require clinical evaluation. Participants were thus classified as fulfilling the symptom-based component of the 2010 ACR criteria for FMS rather than a formal clinical diagnosis of FMS.

The PainDETECT Questionnaire (PDQ) was used to identify participants with suspected presence of neuropathic pain features.^33,34^ The PDQ consists of nine items that measure aspects of sensory quality of pain, the temporal pattern of pain and whether pain radiates. The total score is derived by summing all items with cut-off scores of: ≤12 for unlikely neuropathic pain component, and ≥19 for likely neuropathic component. Total scores between 13 and 18 suggest possible, but uncertain, presence of neuropathic pain.

### Data analysis

Statistical analysis was conducted in RStudio (version 2024.12.1). Participants were categorised into groups based on self-reported CMP at T0: LC without CMP group if reporting no CMP either pre- or post-COVID; LC new-onset CMP group if reporting post-COVID CMP but no pre-COVID CMP; and LC pre-existing CMP group if reporting pre-COVID and post-COVID CMP. Descriptive statistics are presented as mean and standard deviation for continuous variables, or frequency and percentage for categorical variables. Data were screened for potential outliers in questionnaire scores, defined as values exceeding three standard deviations from the sample mean. Identified cases were inspected individually to identify possible data entry errors or inconsistent questionnaire responding. No evidence of data entry errors or response inconsistencies was identified, and all participants were therefore retained for analysis. Incomplete questionnaire responses were coded as missing for the corresponding questionnaire and assessment timepoint. Missing questionnaire data were not imputed. Mixed-effect models were used allowing all available observations from participants with incomplete follow-up data to be included in longitudinal analyses.

Multivariable mixed-effect logistic regression models were fitted to compare odds of meeting ACR criteria for FMS and PDQ classification of likelihood of neuropathic pain component between groups. Group, timepoint, and group x timepoint interaction were modelled as fixed effects, with participant included as random intercept to account for repeated measurements. For PDQ, the outcome was treated as an ordinal variable (unlikely < possible/uncertain < likely) representing increased likelihood of neuropathic component. Where appropriate, post hoc comparisons were estimated using marginal means with adjustment for multiple comparisons using the Holm-Bonferroni method. The significance level was set to 0.05 for all statistical analysis. Sensitivity analyses were conducted adjusting the models for age and sex, with the results reported in the Supplementary Materials.

## Results

### Sample and patient characteristics

A total of 240 participants were included in the study at T0, of which 217 completed T1 and 222 completed T2 with retention rates of 90.4% and 92.5%, respectively (see Figure 1 for detailed participant flow diagram). Only ten (4.2%) individuals reported no CMP either pre-COVID or post-COVID at any follow-up timepoint, and were therefore excluded from analysis for the study to prevent substantial imbalance in subgroup sizes and unstable estimates. Of the initial 240 participants, 131 (100 female) presented with new-onset CMP and 94 (87 female) presented pre-existing CMP before COVID-19 infection in addition to current LC-CMP. The mean age was 47.4 (±11.0) and 51.6 (±11.8) years for the LC new-onset CMP group and the LC pre-existing CMP group, respectively. Table 1 summarises participant demographics, clinical characteristics and LC-associated outcomes for each group. Demographic characteristics of participants who did not complete all follow-up assessments are presented in Supplementary Table S1.

**Figure 1.**
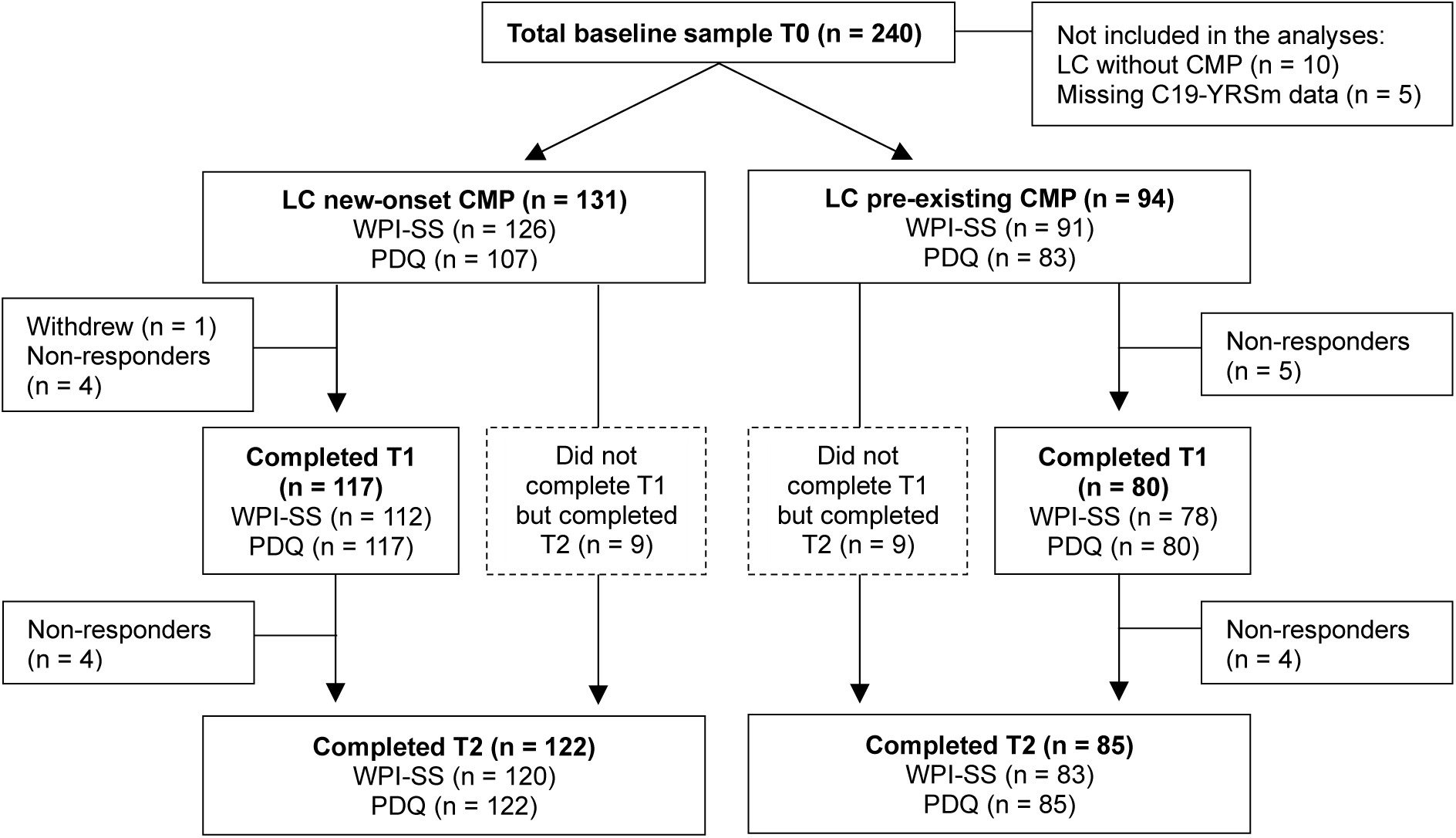
Flow diagram showing participant recruitment, classification at baseline, retention, and questionnaire completion across follow-up period. Participants were classified as having Long COVID without chronic musculoskeletal pain (LC without CMP), Long COVID new-onset chronic musculoskeletal pain (LC new-onset CMP) or Long COVID with pre-existing chronic musculoskeletal pain (LC pre-existing CMP). Numbers indicate participants in each group, follow-up responders and non-responders at each assessment timepoint (baseline = T0, 6-month follow up = T1, 12-month follow-up = T2) and the number of completed questionnaires for each outcome measure at each timepoint (Widespread Pain Index and Symptom Severity scale: WPI-SS; PainDetect Questionnaire: PDQ).

**Table 1.** Summary of demographics, COVID-19 related information and outcomes of Modified COVID-19 Yorkshire Rehabilitation Scale (C19-YRSm) for participants with Long COVID new-onset chronic musculoskeletal pain (LC new-onset CMP) and those with pre-existing chronic musculoskeletal pain prior to COVID-19 infection (LC pre-existing CMP).

| Variables | LC new-onset CMP group<br>(n=131) | LC pre-existing CMP<br>group (n=94) |
| --- | --- | --- |
| **Demographic characteristics** |  |  |
| Age, mean (SD), years | 47.4 (11.0) | 51.6 (11.8) |
| Sex, n (%) |  |  |
| female | 100 (76.3) | 87 (92.6) |
| male | 31 (23.7) | 7 (7.4) |
| Ethnicity, n (%) |  |  |
| Asian (includes any Asian background, e.g. Bangladeshi, Chinese, Indian, Pakistani) | 2 (1.5) | 3 (3.2) |
| Black, African, Black British or Caribbean (includes any black background) | 2 (1.5) | 2 (2.1) |
| Mixed or multiple ethnic groups (includes any mixed background) | 4 (3.1) | 2 (2.1) |
| White (includes any white background) | 123 (93.9) | 87 (92.6) |
| Body mass index, mean (SD), kg/m<sup>2</sup> | 29.6 (13.6) | 30.1 (8.0) |
| Smoking status, n (%) |  |  |
| current occasional smoker | 6 (4.6) | 3 (3.2) |
| current regular smoker | 1 (0.8) | 5 (5.3) |
| ex-smoker | 33 (25.2) | 34 (36.2) |
| never smoked | 91 (69.5) | 52 (55.3) |
| Comorbidities (pre-existing before acute COVID-19 infection), n (%) |  |  |
| type-1 or type-2 diabetes mellitus | 4 (3.1) | 7 (7.4) |
| hypertension | 0 (0.0) | 0 (0.0) |
| autoimmune diseases | 7 (5.3) | 8 (8.5) |
| chronic respiratory conditions | 26 (19.8) | 29 (30.9) |
| chronic heart disease | 0 (0.0) | 2 (2.1) |
| chronic neurological conditions | 0 (0.0) | 3 (3.2) |
| chronic musculoskeletal conditions | 6 (4.6) | 26 (27.7) |
| chronic kidney disease | 0 (0.0) | 3 (3.2) |
| chronic liver disease | 0 (0.0) | 0 (0.0) |
| no pre-existing medical conditions | 63 (48.1) | 23 (24.5) |
| other | 40 (30.5) | 43 (45.7) |
| **COVID-19 related information** |  |  |
| Number of COVID-19 infections, n (%) |  |  |
| 1 infection | 38 (29.0) | 41 (43.6) |
| 2 infections | 57 (43.5) | 33 (35.1) |
| 3 infections | 22 (16.8) | 13 (13.8) |
| 4 infections | 14 (10.7) | 5 (5.3) |
| Infection leading to long-term symptoms, n (%) | (1 missing) | (2 missing) |
| not sure | 4 (3.1) | 3 (3.2) |
| first | 104 (79.4) | 79 (84.0) |
| second | 16 (12.2) | 10 (10.6) |
| third | 5 (3.8) | 2 (2.1) |
| other later COVID-19 reinfection | 1 (0.8) | 0 (0.0) |
| Duration of LC symptoms at T0, mean (SD), months | 32 (13.5) | 32 (13.1) |
| Number of COVID-19 vaccines, n (%) |  | (1 missing) |
| zero | 4 (3.1) | 2 (2.1) |
| one | 1 (0.8) | 3 (3.2) |
| two | 17 (13.0) | 15 (16.0) |
| three | 51 (38.9) | 21 (22.3) |
| four | 58 (44.3) | 52 (55.3) |
| **C19-YRSm outcomes** |  |  |
| Score, mean (SD) |  |  |
| symptom severity | 21.12 (4.67) | 21.94 (4.85) |
| functional disability | 10.41 (3.28) | 10.47 (3.44) |
| overall health | 3.40 (1.80) | 3.94 (1.81) |

Data were collected on average 2.64 (±1.12) years after infection that triggered LC symptoms for T0 (range: 0.10–4.57 years), 3.17 (±1.10) years for T1 (range: 0.59–5.00 years) and 3.69 (±1.09) years for T2 (range: 1.08–5.48 years).

### Fulfilment of symptom-based component of the 2010 ACR diagnostic criteria for FMS at baseline

One participant met the statistical criterion for a potential outlier on WPI-SS scores. Inspection of the questionnaire responses indicated internally consistent responding, suggesting genuine variation within the sample rather than a data entry error. Analyses repeated with and without this participant yielded the same overall pattern of results. Therefore, results are presented for the full sample.

A substantial proportion of participants presented an FMS-like phenotype at T0 based on the 2010 ACR diagnostic criteria for FMS. The highest number was observed in the LC new-onset CMP group, with over three-quarters of individuals meeting the symptom-based ACR criteria for FMS at T0 (n = 99, 78.6%) (Figure 2). In the LC pre-existing CMP group, 64 (70.3%) participants satisfied the symptom-based ACR criteria for FMS at T0. The average WPI score among participants meeting symptom-based ACR criteria for FMS was 10.20 (±4.25) for the LC new-onset CMP group and 10.20 (±3.82) for the LC pre-existing CMP group, indicating a widespread distribution of pain (Table 2). Participants scored on average 9.41 (±1.56) and 8.95 (±1.79) of 12 in the SS scale in the LC new-onset CMP and LC pre-existing CMP groups, respectively, indicating a high level of comorbid symptom severity in both groups.

**Figure 2.**
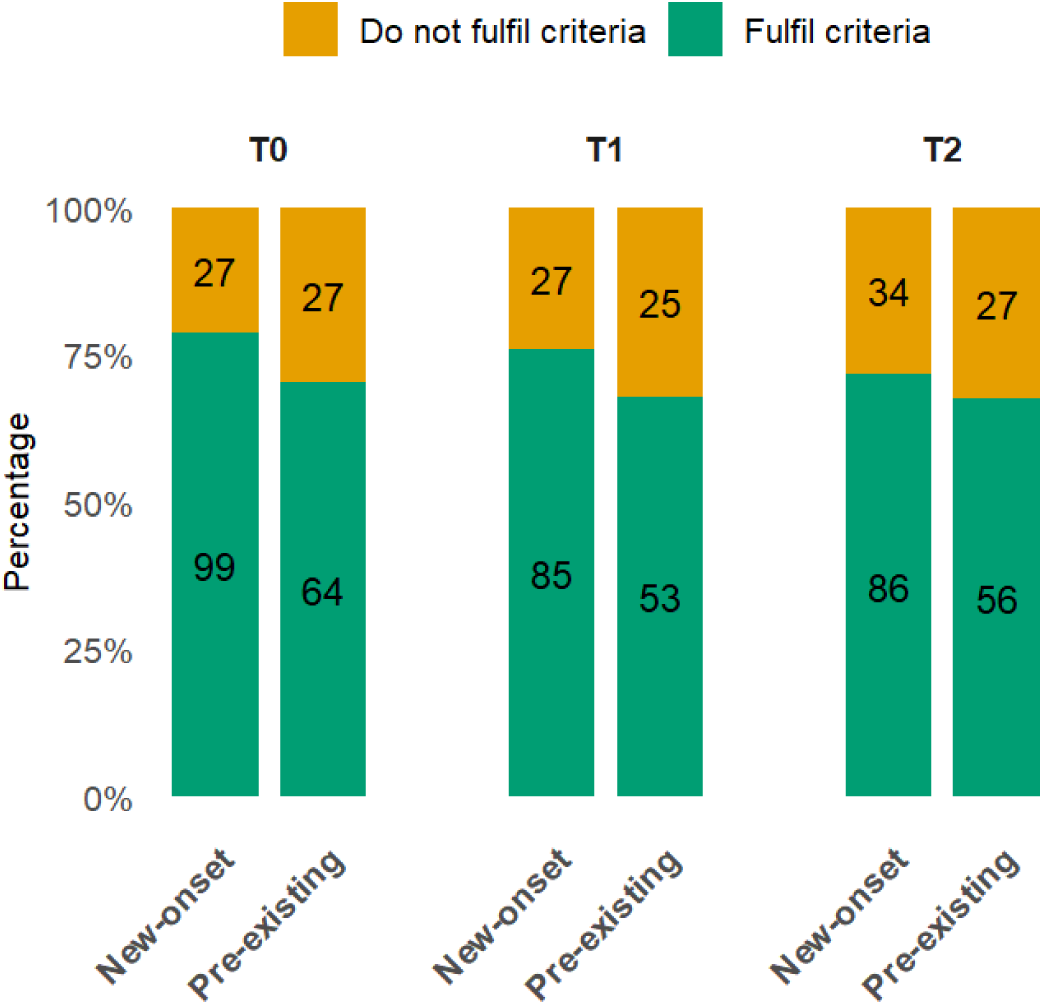
Proportional stacked bar plot showing results of categorisation of individuals with Long COVID (LC) based on fulfilment of the symptom-based component of the 2010 ACR diagnostic criteria for FMS. Proportions are shown in separate bars for the LC new-onset chronic musculoskeletal pain (New-onset) group and for the LC pre-existing chronic musculoskeletal pain (Pre-existing) group, as well as for each follow-up assessment timepoint (T0, T1, T2) approximately six months apart. The numbers inside the stacked bars represent the number of participants in each category.

**Table 2.** Summary of Widespread Pain Index (WPI), Symptom Severity (SS) scale and PainDETECT Questionnaire (PDQ) scores for individuals with Long COVID (LC) reporting new-onset chronic musculoskeletal pain (LC new-onset CMP group) and those that presented pre-existing chronic musculoskeletal pain prior to COVID-19 infection (LC pre-existing CMP group). Results are shown for overall group and for subset of participants that satisfied 2010 ACR diagnostic criteria for FMS based on WPI and SS scales.

| Variables | LC new-onset CMP group |  |  | LC pre-existing CMP group |  |  |
| --- | --- | --- | --- | --- | --- | --- |
| **Overall group** | T0 (n=131) | T1 (n=117) | T2 (n=122) | T0 (n=94) | T1 (n=80) | T2 (n=84) |
| WPI-SS, mean (SD) |  |  |  |  |  |  |
| WPI score | 8.85 (4.58) | 8.29 (4.57) | 7.93 (4.72) | 8.11 (4.65) | 7.64 (4.36) | 8.31 (4.56) |
| SS score | 8.73 (2.00)<br>(5 missing) | 8.44 (2.19)<br>(5 missing) | 8.32 (2.23)<br>(2 missing) | 8.26 (2.00)<br>(3 missing) | 8.17 (2.16)<br>(2 missing) | 8.13 (2.22)<br>(2 missing) |
| PDQ |  |  |  |  |  |  |
| Total score, mean (SD) | 12.9 (8.22) | 12.8 (8.22) | 13.0 (8.60) | 15.6 (7.57) | 14.4 (8.27) | 14.5 (8.11) |
| Likelihood of neuropathic pain based on PDQ, n (%) |  |  |  |  |  |  |
| unlikely | 60 (56.1) | 58 (49.6) | 61 (50.0) | 30 (36.1) | 37 (46.2) | 40 (47.1) |
| possible/uncertain | 21 (19.6) | 33 (28.1) | 33 (27.0) | 23 (27.7) | 17 (21.2) | 20 (23.5) |
| likely | 26 (24.3)<br>(24 missing) | 26 (22.2) | 28 (23.0) | 30 (36.1)<br>(11 missing) | 26 (32.5) | 25 (29.4) |
| **Subset of participants that fulfilled symptom-based component of the 2010 ACR criteria for FMS** | T0 (n=99) | T1 (n=85) | T2 (n=86) | T0 (n=64) | T1 (n=53) | T2 (n=56) |
| WPI-SS, mean (SD) |  |  |  |  |  |  |
| WPI score | 10.2 (4.25) | 9.87 (4.02) | 9.99 (3.78) | 10.2 (3.82) | 9.55 (3.76) | 10.5 (3.73) |
| SS score | 9.41 (1.56) | 9.35 (1.62) | 9.28 (1.51) | 8.95 (1.79) | 9.11 (1.73) | 8.98 (1.90) |
| PDQ |  |  |  |  |  |  |
| Total score, mean (SD) | 14.1 (8.07) | 14.4 (8.18) | 15.5 (8.61) | 17.6 (6.84) | 15.2 (7.22) | 16.0 (7.93) |
| Likelihood of neuropathic pain based on PDQ, n (%) |  |  |  |  |  |  |
| unlikely | 40 (48.2) | 35 (41.2) | 29 (33.7) | 14 (25.9) | 21 (39.6) | 22 (39.3) |
| possible/uncertain | 21 (25.3) | 26 (30.6) | 30 (34.9) | 16 (29.6) | 14 (26.4) | 16 (28.6) |
| likely | 22 (26.5)<br>(16 missing) | 24 (28.2) | 27 (31.4) | 24 (44.4)<br>(10 missing) | 18 (34.0) | 18 (32.1) |

### Longitudinal stability of fulfilment of the symptom-based component of the 2010 ACR criteria for FMS over 12 months

The proportion of individuals fulfilling symptom-based ACR criteria for FMS remained high following the 12-month follow-up period, with more than two thirds of participants in both groups presenting with an FMS-like phenotype at T2. Although the rates of fulfilment at the group level were lower at each follow-up, decreasing from 78.6% to 71.7% at T2 in the LC new-onset CMP group and from 70.3% to 67.5% in the LC pre-existing CMP group, mixed-effects modelling revealed no significant main effect of time (χ²(2) = 3.11, p = 0.211).

Among those with complete follow-up data, 64 of 105 (61.0%) individuals with LC new-onset CMP and 37 of 74 (50.0%) with LC pre-existing CMP fulfilled the symptom-based ACR criteria for FMS at all assessment timepoints (Figure 3). A small proportion of participants remained below the threshold, with 12 (11.4%) individuals in the LC new-onset CMP group and 11 (14.9%) in the LC pre-existing CMP group not fulfilling criteria at any assessment timepoint. The remaining 29 (27.6%) participants with new-onset CMP and 26 (35.1%) with pre-existing CMP moved between fulfilling and not fulfilling symptom-based criteria for CFMS at least once during the 12-month follow-up period.

**Figure 3.**
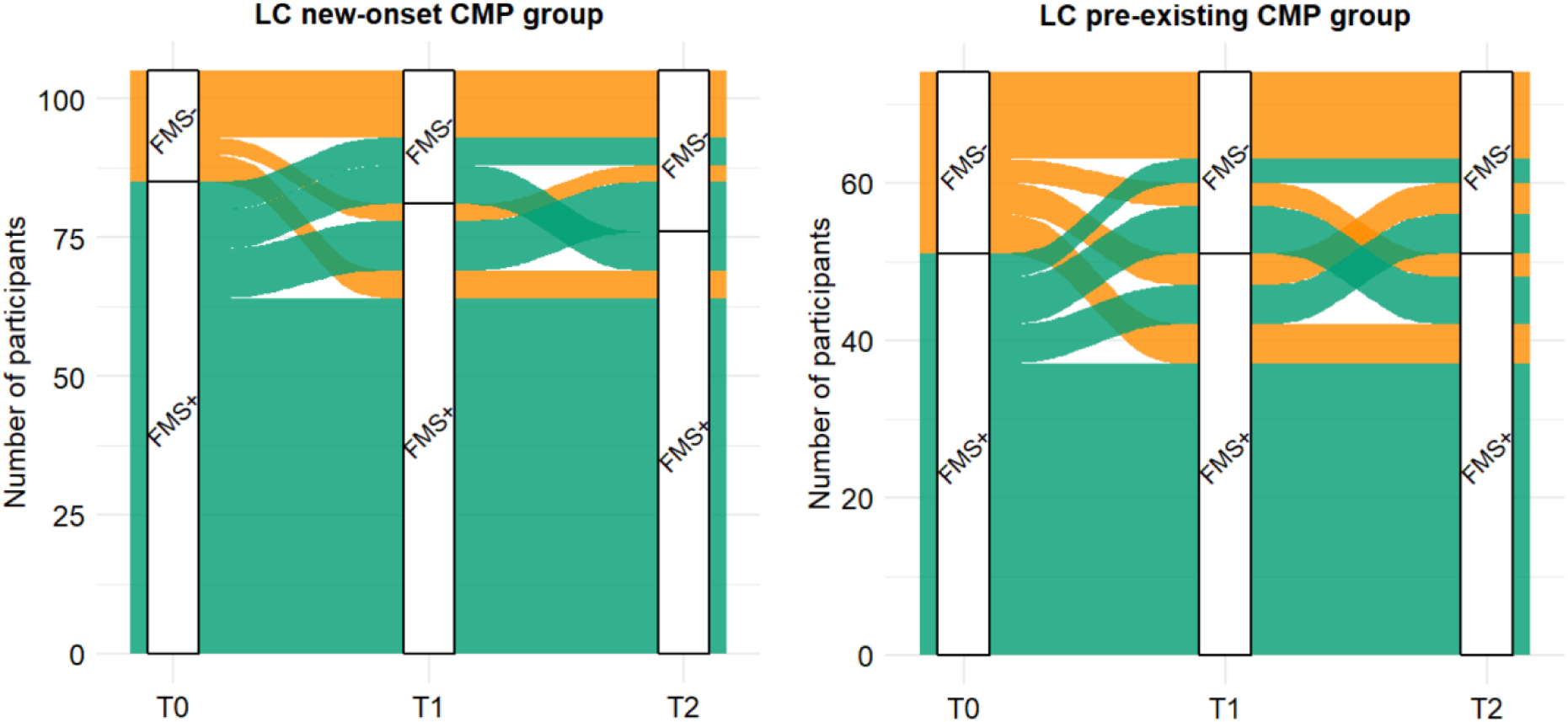
Trajectories of fulfilment of the 2010 ACR symptom-based criteria for Fibromyalgia Syndrome (FMS) over a 12-month follow up period. Alluvial plots showing individual transitions in FMS status between baseline (T0), 6-month (T1) and 12-month (T2) assessment timepoints for the Long COVID new-onset chronic musculoskeletal pain (LC new-onset CMP) group (left) and the Long COVID pre-existing chronic musculoskeletal pain (LC pre-existing CMP) group (right). FMS+ indicates fulfilment of the symptom-based ACR criteria for FMS and FMS-indicates non-fulfilment of the criteria. The width of each flow is proportional to the number of participants following a given trajectory. Only participants with complete data across all assessment timepoints are included in these plots.

### Comparison of FMS symptom-based criteria fulfilment between LC new-onset CMP and pre-existing CMP groups

A mixed-effects binomial logistic regression was conducted to explore whether FMS status differed by group at each timepoint. No evidence of a significant main effect of group (χ²(1) = 1.97, p = 0.160, OR = 0.43, 95% CI [0.14 1.39]) or group x timepoint interaction (χ²(2) = 0.81, p = 0.668) was observed, suggesting no differences between groups in the likelihood of meeting ACR diagnostic criteria for FMS.

### Presence of neuropathic pain features based on the PainDETECT Questionnaire

The mean total PDQ scores ranged from 12.8 to 13.0 for LC new-onset CMP group and 14.4 to 15.6 for the LC pre-existing CMP group across the three follow-up assessments (Table 2). At the group level, 26 participants in the LC new-onset CMP group showed signs of likely neuropathic pain at T0 (24.3%) and T1 (22.2%), going up to 28 (23.0%) at T2. This contrasts with the LC pre-existing CMP group, with over a third of individuals classified as having likely neuropathic pain at T0 (n = 30, 36.1%), followed by 32.5% (n = 26) and 29.4% (n = 25) at subsequent follow-up assessments. Among those with complete follow-up data, 12 of 94 (12.8%) individuals with LC new-onset CMP and 14 of 70 (20.0%) with LC pre-existing CMP were classified as having likely neuropathic pain at all assessment timepoints. Thirty-five (37.2%) individuals in the LC new-onset CMP group and 16 (22.9%) in the LC pre-existing CMP group showed unlikely neuropathic pain across all timepoints. The remaining 41 (43.6%) participants in the new-onset CMP group and 38 (54.3%) with pre-existing CMP changed category of likelihood of neuropathic pain at least once during the 12-month follow-up period.

The mixed-effects ordinal regression model revealed a significant main effect of group (χ²(1) = 9.22, p = 0.002). Specifically, participants in the LC pre-existing CMP group had higher odds of being in higher neuropathic pain likelihood categories based on PDQ scores compared to LC new-onset CMP group (OR = 3.17, 95% CI [1.22 8.18], p = 0.017). There was no evidence of the observed group difference changing over time (group x timepoint interaction effect: χ²(2) = 3.97, p = 0.138).

### Presence of neuropathic pain features in participants that fulfil symptom-based component of the 2010 ACR criteria for FMS

Of those meeting the symptom-based ACR criteria for FMS in the LC new-onset CMP group, 22 (26.5%) were classified as having likely neuropathic pain at T0, with numbers increasing at each follow-up going up to 31.4% (n = 27) at T2. At T0, 48.2% of participants scored 12 or less in the PDQ indicating that a neuropathic component to their pain was unlikely, with the percentage going down to 41.2% and 33.7% in subsequent follow-up assessments.

Across the three assessment timepoints, at least 32.1% of individuals that fulfilled FMS diagnostic criteria in the LC pre-existing CMP group showed signs of likely neuropathic component to their pain, with highest proportion observed at T0 (n = 24, 44.4%). There were 14 (25.9%), 21 (39.6%) and 22 (39.3%) participants classified as unlikely neuropathic pain at T0, T1, and T2, respectively.

## Discussion

This study investigated the persistence of the clinical overlap between LC-CMP and FMS several years after acute COVID-19 infection and whether phenotypic characteristics differ according to pre-existing CMP. Previous studies reported increased rates of FMS cross-sectionally in individuals with LC and up to two years after acute COVID-19 infection. The present findings extend previous cross-sectional observations by showing that the clinical overlap between LC-CMP and FMS persisted over time, remaining evident throughout a 12-month follow-up period, with over two thirds of individuals with LC-CMP exhibiting a symptom profile characteristic of FMS at a mean of two to four years after the triggering infection. The proportion of individuals meeting the symptom-based component of the 2010 ACR criteria for FMS was similar between those reporting LC new-onset CMP and those that presented with pre-existing CMP prior to COVID-19 infection. Furthermore, we report that 20 to 30% of participants demonstrated likely contribution of neuropathic component to their pain, with higher odds in the LC pre-existing CMP group compared to the LC new-onset CMP group.

In our study, over half of the participants with LC-CMP and complete follow-up data continued to fulfil the symptom-based ACR criteria for FMS at all assessment timepoints over the 12-month follow-up period. The high prevalence of an FMS-like phenotype several years after the triggering infection, together with its persistence over a further 12 months, suggests many individuals may have transitioned to a persistent long-term pain state. Across all timepoints, approximately 67.5–78.6% of participants fulfilled the symptom-based ACR criteria for FMS, which is consistent with the estimates observed by Khoja et al.^20^, but higher than previously reported in other cross-sectional studies.^15–17^ This may reflect differences in study populations, as our cohort comprised individuals with self-reported long-term post-COVID symptoms rather than a general post-COVID population, and the present analysis was restricted to participants reporting CMP. Notably, only 10 of the 240 participants (4.2%) reported no pain and were therefore excluded from the analysis to prevent substantial imbalance in subgroup sizes and unstable estimates. The small proportion of pain-free individuals further emphasises the burden of persistent pain within this cohort, even two to four years after acute COVID-19 infection. Together, these findings highlight that LC-CMP should not be regarded as a transient post-viral manifestation expected to resolve spontaneously, but rather as a long-term condition that requires appropriate recognition and ongoing management. Assessment of widespread pain and associated FMS-like symptoms in individuals with LC may be clinically useful, as it could facilitate integration between LC services and established chronic pain rehabilitation pathways. The persistence of LC-CMP symptoms observed in this cohort suggests that rehabilitation approaches that prioritise not only symptom reduction but also the maintenance of physical function, quality of life and participation in valued activities, employment and social roles may be particularly important. ^35^

Notably, we found no significant differences in the proportion of individuals fulfilling symptom-based ACR criteria for FMS between those with new-onset CMP and those with pre-existing CMP. This suggests that LC-CMP presents with a similar profile of FMS-like symptoms regardless of CMP history prior to COVID-19 infection. Although the present study cannot establish a direct link between SARS-CoV-2 infection and the development of an FMS-like phenotype, the substantial and persistent overlap observed across groups may be considered alongside a growing body of literature suggesting a potential role of post-infectious processes in the development and maintenance of FMS-like symptoms.^10,24^ These findings also highlight the potential value of LC as a framework in which to further investigate the mechanisms underlying post-infectious chronic pain syndromes and the development of FMS-like symptoms. Several plausible mechanisms have been proposed. For instance, SARS-CoV-2 neuroinvasion of the central nervous system through binding to nervous system cells expressing the angiotensin-converting enzyme 2 (ACE2) receptor may have direct effects on neuronal and glial function.^36^ Furthermore, the exaggerated immune response to acute SARS-CoV-2 infection and persistent systemic inflammation have been suggested to promote sensitisation of both peripheral and central pain pathways, potentially contributing to amplification of pain processing and development of chronic pain in previously unaffected individuals, or the further amplification or maintenance of sensitisation mechanisms in individuals with prior pain conditions.^9,37^

Up to one third of participants in our cohort reported neuropathic pain features according to the PDQ. This finding is consistent with previous studies showing likely neuropathic pain component in approximately 25% of individuals with LC-CMP using validated self-assessment tools such as the PDQ and the Self-Reported Leeds Assessment of Neuropathic Symptoms and Signs (S-LANNS).^27–29^ While the widespread pain, sensory hypersensitivity and other neurological manifestations commonly observed in LC are largely consistent with nociplastic pain mechanisms and central sensitisation, it is possible that a subset of patients experiences pain secondary to damage or dysfunction of the somatosensory system.^28,38,39^ Indeed, evidence of peripheral neuropathy has been reported in some individuals with LC,^38,40,41^ supporting the potential contribution of neuropathic pain mechanisms in at least a subset of cases. Nevertheless, nociplastic and neuropathic pain mechanisms are not mutually exclusive and may coexist along a continuum, with substantial overlap in their clinical presentation. Accordingly, neuropathic-like sensory symptoms such as burning, tingling and numbness may occur as a result of underlying neuropathy or as manifestations of altered sensory processing within a predominantly nociplastic pain syndrome. On that note, comparable rates of neuropathic-like sensory symptoms have been reported in FMS patients,^42–44^ despite the condition being generally regarded as nociplastic in nature, further emphasising the clinical overlap between FMS and LC. In our study, patients reporting pre-existing CMP prior to COVID-19 infection showed higher odds of presenting neuropathic pain features compared to those with new-onset CMP. One possible explanation is that, in this subgroup, the prolonged exposure to pain may have cumulative effects and contribute to enhanced sensitisation of nociceptive pathways through maladaptive neuroplasticity.^45^ Alternative explanations, such as differences in comorbidities or the existence of distinct pain phenotypes, should also be considered. Collectively, these findings reflect the complex heterogenous nature of FMS and LC, reinforcing the importance of individual assessment and the need for a multimodal therapeutic approach tailored to individual patient presentation.

This study has several limitations. First, recruitment through LC support networks and the restriction of analyses to participants reporting CMP may have introduced selection bias and limited generalisability to the wider LC population. Second, assessment of the phenotypic characteristics of LC-CMP relied on self-report assessment tools, which are susceptible to recall and response bias. In addition, information on pain duration was not collected, limiting our ability to fully characterise the temporal nature of MSK pain within this cohort. Furthermore, use of the symptom-based component of the 2010 ACR criteria means a formal diagnosis of FMS could not be established, which would require comprehensive clinical examination. Additionally, there is an overlap between core LC symptoms and domains included in the ACR criteria which may have influenced classification rates. Therefore, the observed symptom overlap between LC-CMP and FMS should be interpreted as evidence of phenotypic similarity rather than proof that these conditions represent the same diagnosis. We applied the 2010 ACR criteria, as these were the most widely used in clinical practice at the time of study design, although revised ACR^46^ criteria released in 2016 are available and may yield different estimates. Third, the sample was predominantly female and middle-aged. The potential contribution of hormonal factors to the observed symptom profile cannot be excluded given that menopausal symptoms overlap with LC and FMS symptom domains. Future studies should consider adjustment for menopausal status and hormonal replacement therapy use to help clarify hormonal influences on symptom burden. Fourth, the study did not include comparator groups, such as individuals with LC without pain, individuals recovered from COVID-19 infection or patients with a clinical diagnosis of FMS, limiting assessment of the specificity of the observed symptom profile. Finally, while the PDQ is a validated neuropathic pain screening tool, alternative instruments are available, such as the S-LANNS and Douleur Neuropathique 4 (DN4),^47,48^ that may yield different neuropathic pain classification and prevalence estimates. Our findings should be interpreted as reflecting suspected presence of neuropathic pain features as identified by the PDQ, not as definitive presence of neuropathic pain. Objective evidence of damage or dysfunction of the somatosensory nervous system is needed to confirm the involvement of neuropathic pain mechanisms, through techniques such as nerve conductions studies, skin biopsy and quantitative sensory testing.

## Conclusion

This study shows that the symptom overlap between LC-CMP and FMS remains evident up to four years after acute COVID-19 infection and persists over a further 12-month follow-up period. These findings suggest that LC-CMP should not be regarded as a transient post-viral manifestation, but as a persistent and clinically significant condition requiring appropriate recognition and long-term management. Many individuals experience a long-term symptom burden characteristic of chronic widespread pain conditions, supporting the need for comprehensive assessment and closer integration with established chronic pain rehabilitation services. The heterogeneity of LC-CMP highlights the need for further phenotypic and mechanistic research to improve understanding of its underlying pathophysiology. Insights gained from LC-CMP research may ultimately improve our understanding of FMS and other post-infectious chronic pain conditions.

## Acknowledgements

The authors would like to express their heartfelt gratitude to all participants for their invaluable contribution to this study. The authors also wish to acknowledge the software development team at ELAROS 24/7 for their support in the development and maintenance of the digital platform used to conduct this research.

## Author contributions

BSP, ID, AJC, CB and MS contributed to the conception and design of the study. BSP collected and analysed the data. BSP drafted the manuscript. All authors contributed to data interpretation, critically revised the manuscript, and approved the version for publication.

## Ethical considerations

Ethical approval for this study was obtained from the University of Leeds’ Faculty of Medicine Ethics committee (MREC 22-019).

## Consent to participate

Electronic informed consent was obtained from all participants prior to study participation.

## Consent for publication

Not applicable

## Data availability statement

All data are available upon reasonable request to the corresponding author.

