## Supplementary for "Persistent Fibromyalgia-like symptom presentation in Long COVID-associated chronic musculoskeletal pain: a longitudinal cohort study"

Manoj Sivan, Leeds Institute of Rheumatic and Musculoskeletal Medicine, 2nd Floor, Chapel Allerton Hospital, Chapeltown Road, Leeds, LS7 4SA

**Table S1. Summary of demographics, COVID-19 related information and outcomes of Modified COVID-19 Yorkshire Rehabilitation Scale (C19-YRSm) for completers and non-completers.** Completers includes participants who completed all assessment timepoints as well as those who did not complete the 6-month follow but completed the 12-month follow up. Non-completers comprises participants who completed only the baseline assessment (n = 9) and those who completed baseline and 6-month follow-up assessments but not the 12-month follow-up (n = 8), with data are combined across groups due to the small sample size.

| **Variables** | **Completers** (n = 208) | **Non-completers** (n = 17) |
|---|---|---|
| **Demographic characteristics** | | |
| Age, mean (SD), years | 49.3 (11.3) | 47.0 (13.4) |
| Sex, n (%) | | |
| female | 173 (83.2) | 14 (82.4) |
| male | 35 (16.8) | 3 (17.6) |
| Ethnicity, n (%) | | |
| Asian (includes any Asian background) | 4 (1.9) | 1 (5.9) |
| Black, African, Black British or Caribbean (includes any black background) | 4 (1.9) | 0 (0.0) |
| Mixed or multiple ethnic groups (includes any mixed background) | 6 (2.9) | 0 (0.0) |
| White (includes any white background) | 194 (93.3) | 16 (94.1) |
| Body mass index, mean (SD), $kg/m^2$ | 29.8 (12.0) | 29.8 (5.1) |
| Smoking status, n (%) | | |
| current occasional smoker | 7 (3.4) | 2 (11.8) |
| current regular smoker | 6 (2.9) | 0 (0.0) |
| ex-smoker | 60 (28.8) | 7 (41.2) |
| never smoked | 135 (64.9) | 8 (47.1) |
| Comorbidities (pre-existing before acute COVID-19 infection), n (%) | | |
| type-1 or type-2 diabetes mellitus | 11 (5.3) | 0 (0.0) |
| hypertension | 0 (0.0) | 0 (0.0) |
| autoimmune diseases | 11 (5.3) | 4 (23.5) |
| chronic respiratory conditions | 49 (23.6) | 6 (35.3) |
| chronic heart disease | 1 (0.5) | 1 (5.9) |
| chronic neurological conditions | 3 (1.4) | 0 (0.0) |
| chronic musculoskeletal conditions | 28 (13.5) | 4 (23.5) |
| chronic kidney disease | 2 (1.0) | 1 (5.9) |
| chronic liver disease | 0 (0.0) | 0 (0.0) |
| no pre-existing medical conditions | 82 (39.4) | 4 (23.5) |
| other | 74 (35.6) | 9 (52.9) |
| **COVID-19 related information** | | |
| Number of COVID-19 infections, n (%) | | |
| 1 infection | 72 (34.6) | 7 (41.2) |
| 2 infections | 85 (40.9) | 5 (29.4) |
| 3 infections | 32 (15.4) | 3 (17.6) |
| 4 infections | 17 (8.2) | 2 (11.0) |
| Infection leading to long-term symptoms, n (%) | | |
| not sure | 6 (2.9) | 1 (5.9) |
| first | 168 (80.8) | 15 (88.2) |
| second | 25 (12.0) | 1 (5.9) |
| third | 7 (3.4) | 0 (0.0) |
| other later COVID-19 reinfection | 1 (0.5) | 0 (0.0) |
| Duration of LC symptoms at T0, mean (SD), months | 32.2 (13.1) | 29.0 (15.2) |
| Number of COVID-19 vaccines, n (%) | | |
| zero | 5 (2.4) | 1 (5.9) |
| one | 4 (1.9) | 0 (0.0) |
| two | 29 (13.9) | 3 (17.6) |
| three | 67 (32.2) | 5 (29.4) |
| four | 102 (49.0) | 8 (47.1) |
| **C19-YRSm outcomes** | | |
| Score, mean (SD) | | |
| symptom severity | 21.4 (4.65) | 21.8 (6.0) |
| functional disability | 10.4 (3.36) | 10.5 (3.2) |
| overall health | 3.6 (1.84) | 3.9 (1.6) |

**Table S2. Sensitivity analysis of fulfilment of symptom-based criteria for Fibromyalgia Syndrome (FMS) adjusting for the effects of age and sex.** A summary of outputs from mixed-effects logistic regression models examining the effects of group (Long COVID new-onset chronic musculoskeletal pain vs Long COVID pre-existing chronic musculoskeletal pain), timepoint (baseline, 6- and 12-month follow-up), and the group × timepoint interaction on odds of meeting the symptom-based component of the 2010 American College of Rheumatology (ACR) criteria for FMS is shown. Participant was included as a random intercept to account for repeated measurements. Values represent Wald $\chi^2$ statistics and associated p values from Type III analyses of variance. Sensitivity analyses adjusting separately for age and sex yielded results consistent with the unadjusted model, with no changes in the statistical significance of observed effects.

| Effect | $\chi^2$(df) | p value |
|---|---|---|
| **Unadjusted model** | | |
| Group | 1.97(1) | 0.160 |
| Survey | 3.11(2) | 0.211 |
| Group x Survey | 0.81(2) | 0.668 |
| **Age-adjusted model** | | |
| Group | 1.99(1) | 0.158 |
| Survey | 3.11(2) | 0.211 |
| Age | 0.02(1) | 0.879 |
| Group x Survey | 0.81(2) | 0.668 |
| **Sex-adjusted model** | | |
| Group | 2.63(1) | 0.105 |
| Survey | 3.18(2) | 0.204 |
| Sex | 1.68(1) | 0.195 |
| Group x Survey | 0.83(2) | 0.661 |

**Table S3. Sensitivity analysis of PainDETECT Questionnaire (PDQ) classification of likelihood of neuropathic pain component adjusting for the effects of age and sex.** A summary of outputs from cumulative link mixed models examining the effects of group (Long COVID new-onset chronic musculoskeletal pain vs Long COVID pre-existing chronic musculoskeletal pain), timepoint (baseline, 6- and 12-month follow-up), and the group × timepoint interaction on PDQ classification is shown. PDQ classification was treated as an ordinal variable (unlikely < possible/uncertain < likely) representing increased likelihood of neuropathic component. Participant was included as a random intercept to account for repeated measurements. Values represent Wald $\chi^2$ statistics and associated p values from Type III analyses of variance. Sensitivity analyses adjusting separately for age and sex yielded results consistent with the unadjusted model, with no changes in the statistical significance of observed effects.

| Effect | $\chi^2$(df) | p value |
|---|---|---|
| **Unadjusted model** | | |
| Group | 9.22(1) | 0.002** |
| Survey | 2.65(2) | 0.266 |
| Group x Survey | 3.96(2) | 0.138 |
| **Age-adjusted model** | | |
| Group | 9.13(1) | 0.002** |
| Survey | 2.65(2) | 0.266 |
| Age | 0.01(1) | 0.910 |
| Group x Survey | 3.97(2) | 0.138 |
| **Sex-adjusted model** | | |
| Group | 7.45(1) | 0.006** |
| Survey | 2.66(2) | 0.265 |
| Sex | 3.38(1) | 0.066 |
| Group x Survey | 4.05(2) | 0.132 |
